# Vulnerability and burden of all-cause mortality associated with particulate air pollution increased during COVID-19 pandemic: a nationwide observed study in Italy

**DOI:** 10.1101/2020.10.02.20206052

**Authors:** Tingting Ye, Rongbin Xu, Wenhua Yu, Zhaoyue Chen, Yuming Guo, Shanshan Li

## Abstract

**Background:** Limited evidence is available on the health effects of particulate matter (i.e. PM_2.5_, particulate matter with an aerodynamic diameter < 2.5μm; PM_10_, < 10μm; PM_2.5-10_, 2.5-10μm) during the pandemic of COVID-19 in Italy.

**Objectives:** To examine the associations between all-cause mortality and daily PM_2.5_, PM_2.5-10_, and PM_10_ in the pandemic period, and compare them to the normal periods (2015-2019) in Italy.

**Methods:** We collected daily data regarding all-cause (stratified by age and gender), and PM_2.5_, PM_2.5-10_, and PM_10_ for 107 Italian provinces from 1, January 2015 to 31, May 2020. A time-stratified case-cross design with the distributed lag non-linear model was used to examine the association between PM and all-cause mortality during the first three months of the COVID-19 outbreak (March to May in 2020) and the same months in 2015-2019. We also compared the counts and fractions of death attributable to PM in two periods.

**Results:** Overall, Italy saw an increase in daily death counts while slight decreases in PM concentrations in 2020 pandemic period compared to same months of 2015-2019. Mortality effects were significant in lag 0-3 days for PM_2.5_, lag 0-2 for PM_10_, and lag 0-1 for PM_2.5-10_. Each 10 µg/m^3^ increase in PM was associated much higher increase in daily all-cause mortality during 2020 pandemic period compared to the same months during 2015-2019 [increased mortality rate: 7.24 % (95%CI: 4.84%, 9.70%) versus 1.69% (95%CI: 1.12%, 2.25%) for PM_2.5_; 3.45 % (95%C: 2.58%, 4.34%) versus 1.11% (95%CI: 0.79%, 1.42%) for PM_10_, 4.25% (95%CI: 2.99%, 5.52%) versus 1.76% (95%CI: 1.14%, 2.38%) for PM_2.5-10_]. The counts and fractions of deaths attributable to PM were higher in 2020 than the normal periods for PM_2.5_ (attributable death counts: 20,062 in 2020 versus 3,927 per year in 2015-2019; attributable fractions: 10.2% versus 2.4%), PM_10_ (15,112 versus 3,999; 7.7% versus 2.5%), and PM_2.5-10_ (7,193 versus 2303; 3.7% versus 1.4%).

**Conclusions:** COVID-19 pandemic increased the vulnerability and excess cases of all-cause mortality associated with short-term exposure to PM_2.5_, PM_2.5-10_ and PM_10_ in Italy, despite a decline in air pollution level. This suggests using historical PM-mortality association to calculate health benefits associated with reduction in PMs has big uncertainties.

## 1. Introduction

Coronavirus Disease 2019 (COVID-19) pandemic caused by the severe acute respiratory syndrome coronavirus 2 (SARS-CoV-2) virus^1^ has substantially affected human society, including healthcare, economic structure, social relationships. The measures and responses to control virus transmission can protect human health, but it also results in unprecedented side-effects. Even though the severe health impacts of the COVID-19 emergence remain the top priority, it is still unknown how the pandemic may affect the association between environmental exposure and health, notably the health impacts of air pollution, particularly for ambient particulate matter (PM) air pollutant which is an important risk factor for cardiovascular and respiratory health outcomes.^2 3^

The COVID-19 pandemic has observed a noteworthy decline in anthropogenic PM in many countries, such as China,^4 5^ Morocco,^6^ Malaysia,^7^ India,^8^ Brazil,^9^ United States,^10^ and Spain^11^ due to the reduced social activities and vehicle exhaust emission.^12 13^ To control the COVID-19 outbreak, Italian government imposed a quarantine, restricting the migrations of the population and additional lockdown restrictions mandating the temporary closure of non-essential shops and businesses first in ten municipalities of the province of Lodi in Lombardy and neighboring municipalities in the northern region at around February 21, 2020, and then in early-March the quarantine measures were expanded to the entire country.^14^ Alicandro et al^15^ suggested that Italy’s first wave of the COVID-19 pandemic has ended in May because no excess mortality but COVID-19 deaths were probably over-registered. During the first wave of COVID-19 pandemic (March to May) in Italy, the severe limitation of people movements determined a significant reduction of PM_10_, PM_2.5_ pollutants concentration mainly due to vehicular traffic, especially over hard-hit northern Italy (i.e. Milan^16^ and Roma^17^). It is expected that reductions in PM could reduce burden of the air pollution-related diseases, if the magnitude of mortality/morbidity risks associated with PM would not change. However, it is still unclear whether this hypothesis is correct, because the exposure-response association could be changed due to the alteration of people’s behaviors and reorganized medical resources during the COVID-19 pandemic.

Previous assessments have focused on the effects of PM on the death of COVID-19.^18-21^ However, the number of COVID-19 deaths in official reports may be significantly lower than the actual death toll due to the insufficient testing availability, especially during rapid growth phases of the epidemic.^22^ In addition, indirect deaths from the pandemic may increase because of the limitation of health care resources, especially for the patients who were scheduled for surgery or treated but suspended due to the pandemic.^23 24^ On the other hand, some susceptible population are concerned about being infected when going to a hospital.^25^

The overlap of vulnerable populations related to COVID-19 and unrelated makes the measurement of the pandemic impacts complexed. Unlike the deaths attributed to COVID-19, all-cause mortality could be an accurate measurement to estimate the direct and indirect effects of the pandemic on deaths.^26^ Therefore, understanding whether the COVID-19 pandemic could switch the association of PM with total all-cause mortality or not would provide important clues regarding health promotion and air pollution control.

In this study, we aimed to quantify the association between short-term exposure to PM and all-cause mortality in Italy during the first wave of COVID-19 pandemic and compare it with that of the same months in 2015-2019 to identify the changes in PM-mortality association.

## 2. Methods

### 2.1 Study setting

Italy is a country consisting of a peninsula delimited by the Alps and surrounded by several islands, covering a total area of 301,340 km^2^. There are currently 107 provinces (second level constituent entities) in Italy, within 20 regions (first-level constituent entities). Its northern regions (Lombardia, Veneto, Piemonte, Emilia Romagna) together host 39% of the national population, and approximately one-half of the Italian GDP is produced there. Such a spatial concentration of economic activities involves the industrial manufacturing sectors to the largest extent, and the consequent high level of emissions is at least in part responsible for heavy pollution in the region.^27^ During the COVID-19 pandemic, Italy has been one of the worst countries affected by the spreading of coronavirus, especially the northern regions.^28^ Here, we performed analyses with daily air quality data from national monitoring stations and daily all-cause mortality data at province level.

### 2.2 Mortality data

We collected daily all-cause mortality from Italian National Institute of Statistics (ISTAT) from January 1, 2015 to May 31, 2020. The mortality data covers 7,357 municipalities in 107 provinces, representing 95.0% of Italy population since January 1, 2015. Daily counts of all-cause mortality were aggregated at province level and stratified by gender and age-specific groups (< 65 years and ≥ 65 years).

### 2.3 Environmental exposure data

Air pollution data were downloaded from European Environment Agency (EEA) air quality database. The database includes hourly PM_2.5_, PM_10_, ozone (O_3_), nitrogen dioxide (NO_2_), sulfur dioxide (SO_2_), and carbon monoxide (CO). The 24-h average concentrations of PM_2.5_, PM_10_, NO_2_, SO_2_, and CO were calculated as daily concentration, while we used the maximum 8-h average concentrations of O_3_ as its daily value. Daily PM_2.5-10_ was calculated as the difference between 24-h average PM_10_ and 24-h average PM_2.5_.^29^ Daily concentrations of air pollutants at station level in the same province were averaged one daily times series. If a province only had one monitoring station, data from this station was used to represent the exposure level of this province.

To allow adjustment for the meteorological factors, we collected the ERA5 hourly surface (at 2 meters above the land surface) ambient temperature and ambient dew point temperature at 0.1°×0.1° spatial resolution from the ERA5-Land hourly data. Hourly data were averaged into daily values. We calibrated the collected temperature data with the observed meteorological data through random forest models (see Supplementary materials for detail), and then we linked the data to the centroid of each municipality based on longitude and latitude. We then calculated daily mean relative humidity from the calibrated ERA5 daily mean temperature and ERA5 daily mean dew point temperature, using the algorithm provided by the “humidity” R package.^30^ Weather data at municipality level were aggregated into province level by averaging observations of all municipalities within the province.

### 2.4 Statistical analysis

A time-stratified case-crossover design was used to examine the association between PM air pollution and all-cause mortality. The design compares the air pollution exposure in the case period when events occurred with air pollution exposures in control periods to compare the differences in exposure which might explain the differences in the daily number of cases. In this study, the cases and controls were matched by day of the week in the same calendar month and the same province, to control for the effects of day of the week, seasonality, long-term trend, and spatial variation on deaths or pollution. The Quasi-Poisson regression^31^ allowing for over-dispersion was applied to perform time-stratified case-crossover design. To determine an appropriate lag time (i.e., the number of days between exposure and the estimated effect) for PM to be used in the main analyses, we compared a variety of lag days and choose all significant lags as the maximum lag day (see Supplementary Figure S1). A linear function was used for PM concentrations while a 3 degrees of freedom natural cubic spline was used for lags. Our initial analyses showed that significant mortality effects were observed in lag 0, 1, 2, and 3 days for PM_2.5_, lag 0, 1, and 2 days for PM_10_, and lag 0, 1 days for PM_2.5-10_. Therefore, we used cumulative effects along lag 0-3 days for PM_2.5_, lag 0-2 for PM_10_, and lag 0-1 for PM_2.5-10_ for subsequent analyses. We have controlled for potential nonlinear and lagged confounding effects of weather conditions, with 3 degrees of freedom natural cubic spline for 21-day moving averages^32 33^ of daily mean temperature and daily mean relative humidity, respectively.

To compare the associations between PM air pollution and all-cause mortality during COVID pandemic and pre-outbreak periods, we performed above analyses for 2020 COVID pandemic period (from March 1 to May 31, 2020) and the same months during 2015-2019, respectively. The analyses for PM_2.5_, PM_10_ and PM_2.5-10_ were performed separately, to avoid their high collinearity. Fixed effect meta-regression was used to compare the magnitude of the mortality risks associated with PM air pollution in different sub-groups.

To estimate the burden of mortality attributable to PM, the attributable number deaths (AD) caused by PM were calculated every day and total AD was generated by summing the AD during the study period.^34^ The corresponding attributable fractions (AF) of mortality were calculated by dividing the total AD by the death toll.

### 2.5 Sensitivity analyses

Sensitivity analyses were performed to examine the robustness of the results. We tested the variation of the PM pollution-mortality association in normal period by replacing the study period of 2015-2019 with every single year. To evaluate potential impacts of gaseous pollutants on the associations between PM air pollution and mortality, we also performed multi-pollutant models through adjusting for different combinations of NO_2_, CO, O_3_, and SO_2_.

All the analyses were performed by the R software (v. 3.6.1). The ‘dlnm’ package was used to perform the distributed lag non-linear models for PM, describing simultaneously the linear relationship along air pollution and non-linear along lags; the ‘gnm’ package was used to perform conditional Poisson regression.^35^ The “mvmeta” package was used to perform meta-regression.^36^ The relative risks (RRs) with 95% confidence intervals (CIs) per 10 µg/m^3^ change in PM concentration were reported. For all statistical tests, a *p* value of 0.05 (two-tailed) was considered statistically significant.

## 3. Results

There were 1,000,459 (51.9% females; 89.4% aged ≥ 65 years) all-cause deaths in the years from March to May during 2015 and 2020. The death counts in the pandemic period were significantly higher than the same months during 2015-2019. For instance, the daily average mortality counts have risen by 24.61% from 837 ± 62 in 2015-2019 to 1043 ± 325 in 2020 for men and by 19.98% for women from 911 ± 80 to 1093 ± 271 (Table 1). There were no significant trends in mean temperature and relative humidity from 2015 to 2020, but the pandemic period witnessed a slight decrease in relative humidity versus an increase in mean temperature compared with the same months during 2015-2019 (Table 1).

**Table 1.** Descriptive statistics for temperature, relative humidity, and death counts during the first three months of COVID-19 pandemic in 2020 and the same months in 2015-2019.

|  | 2015 | 2016 | 2017 | 2018 | 2019 | 2015-2019 | 2020 | *p* value |
| --- | --- | --- | --- | --- | --- | --- | --- | --- |
| Temperature (°C) | 12.32 (4.51) | 12.20 (4.14) | 13.01 (4.07) | 12.75 (5.18) | 11.48 (3.1) | 12.35 (4.29) | 12.52 (4.40) | < 0.001 |
| RH (%) | 67.62 (11.45) | 69.12 (10.43) | 65.64 (10.96) | 72.76 (9.43) | 68.85 (12.07) | 68.80 (11.15) | 65.41 (12.08) | < 0.001 |
| PM<sub>2.5</sub> (µg/m<sup>3</sup>) | 15.60 (10.23) | 12.60 (7.71) | 13.59 (9.50) | 13.38 (8.49) | 11.77 (8.02) | 13.55 (9.01) | 12.52 (7.57) | < 0.001 |
| PM<sub>10</sub> (µg/m<sup>3</sup>) | 23.05 (12.25) | 20.42 (13.46) | 20.73 (12.45) | 22.66 (12.21) | 19.71 (12.95) | 21.40 (12.7) | 20.54 (13.95) | < 0.001 |
| PM<sub>2.5-10</sub> (µg/m<sup>3</sup>) | 8.86 (5.83) | 8.24 (6.78) | 8.46 (5.66) | 8.48 (6.16) | 8.49 (6.07) | 8.51 (6.83) | 8.14 (7.43) | < 0.001 |
| Daily Average deaths counts |  |  |  |  |  |  |  |  |
| Age < 65 years | 200 (17) | 192 (15) | 190 (15) | 192 (17) | 187(17) | 192 (17) | 196 (37) | 0.097 |
| Age ≥ 65 years | 1590 (150) | 1523 (104) | 1544 (82) | 1548 (154) | 1573 (125) | 1555 (128) | 1939 (556) | < 0.001 |
| Female | 941 (93) | 887 (66) | 903 (53) | 906 (93) | 918 (80) | 911 (80) | 1093 (271) | < 0.001 |
| Male | 849 (70) | 828 (50) | 831 (46) | 835 (73) | 842 (64) | 837 (62) | 1043 (325) | < 0.001 |
| Total | 1790 (157) | 1715 (109) | 1734 (88) | 1740 (161) | 1759 (135) | 1748 (135) | 2136 (590) | < 0.001 |

Note: Mean and SD (Standard deviation) were presented; p-Value was calculated by independent Two-Sample t-Test comparing observations in 2020 versus all observations in 2015-2019.

There were slight reductions in PM_10_, PM_2.5_ and PM_2.5-10_ concentrations over 2015-2020 (Table 1). The average (± SD) PM_2.5_ concentration during March and May reduced from 15.60 ± 10.23 μg/m^3^ in 2015 to 12.52 ± 7.57 μg/m^3^ in 2020, PM_10_ from 23.05 ± 12.25 μg/m^3^ to 20.54 ± 13.95 μg/m^3^, and PM_2.5-10_ from 8.86 ± 5.83 μg/m^3^ to 8.14 ± 7.43 μg/m^3^,with all the differences being statistically significant (p < 0.001).

Figure 1 shows the spatial variation in all-cause deaths, PM_2.5_, PM_10_ and PM_2.5-10_ during March and May in 2020 and 2015-2019, respectively. The figure presented the difference by subtracting the average of 2015-2019 from the daily value in 2020. The northern region had increased number of deaths in 2020. The PM concentrations declined slightly from 2015-2019 to 2020.

**Fig. 1:**
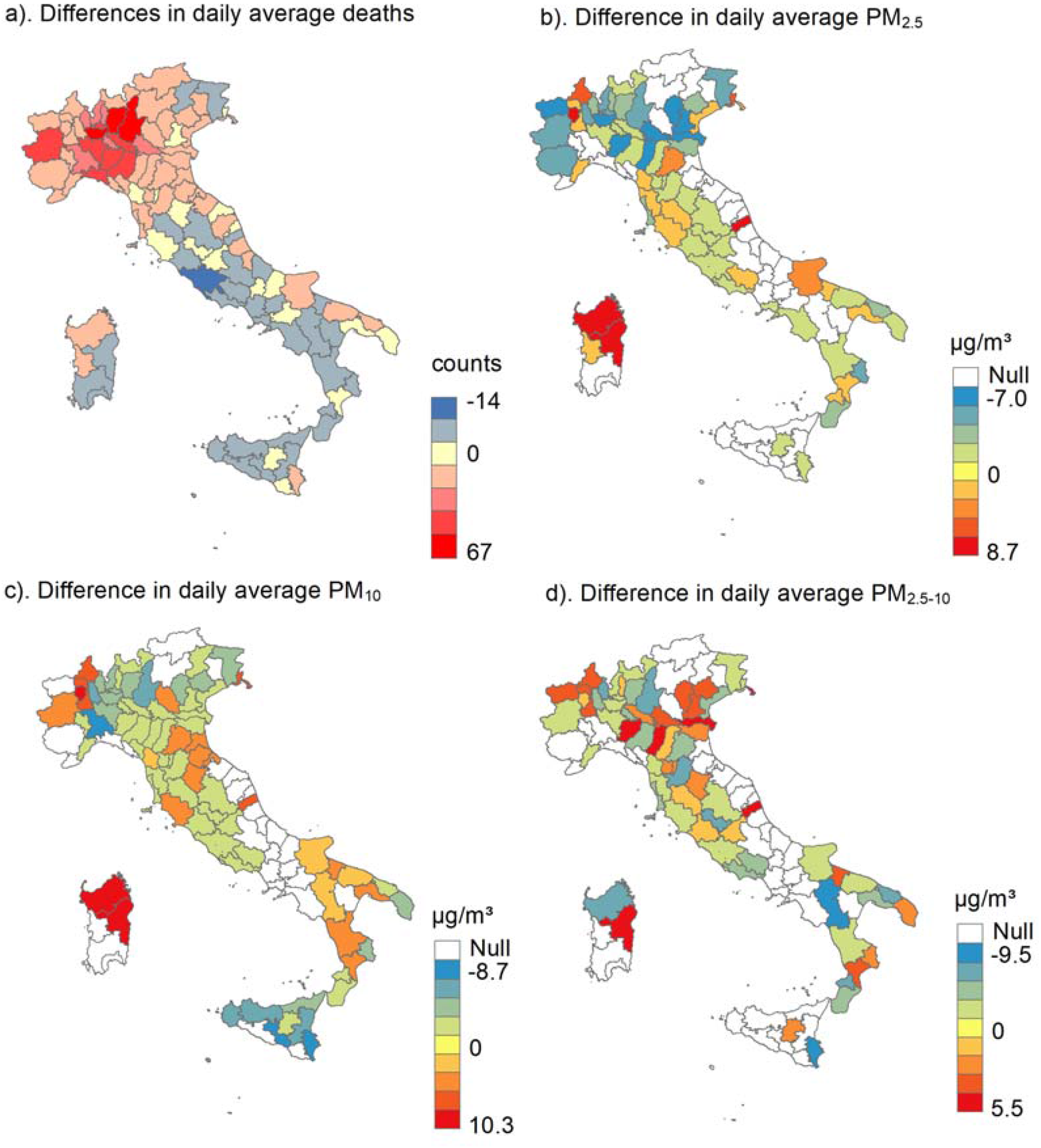
Difference in daily average deaths and the air pollutants in 107 Italy provinces during March and May.

Cumulative RRs along lag 0-3 days for PM_2.5_, lag 0-2 for PM_10_, and lag 0-1 for PM_2.5-10_ for all-cause mortality and group-specific mortality are shown in Figure 2. Each 10 µg/m^3^ increase in PM was associated much higher increase in daily all-cause mortality during 2020 pandemic period compared to the same months during 2015-2019 [increased mortality risk: 7.24 % (95%CI: 4.84, 9.70) versus 1.69% (95%CI: 1.12, 2.25) for PM_2.5_; 3.45 % (95%C: 2.58, 4.34) versus 1.11% (95%CI: 0.79, 1.42) for PM_10_, 4.25 % (95%CI: 2.99, 5.52) versus 1.76% (95%CI: 1.14, 2.38) for PM_2.5-10_]. Such disparity in the PM-mortality associations were consistent among different gender and age groups.

**Fig. 2:**
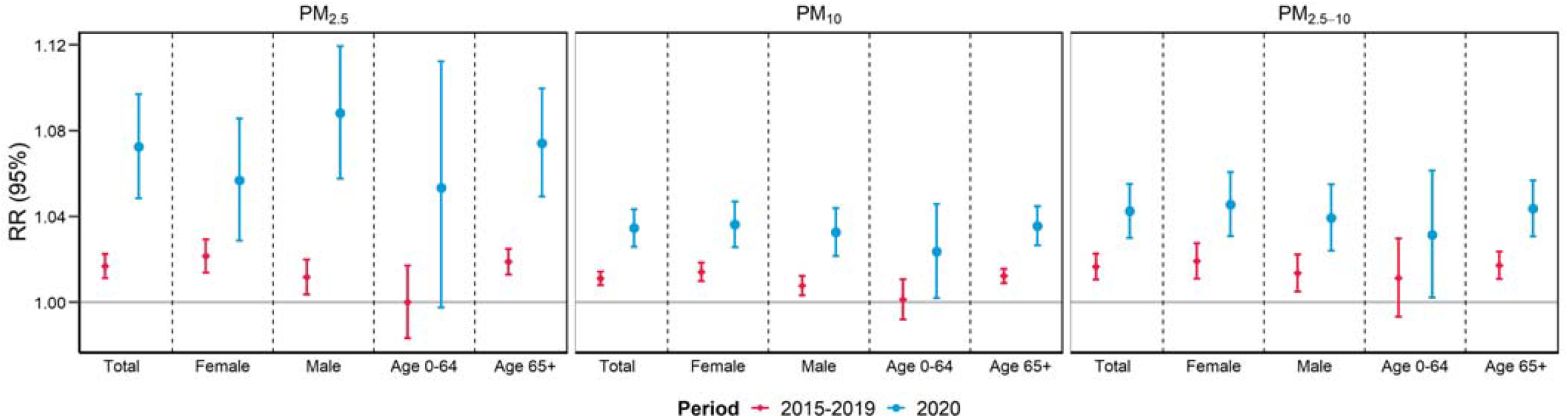
Cumulated RR of all-cause mortality and group-specific mortality associated with a 10 µg/m^3^ increase in the concentrations of PM air pollutants. P-value for difference (2020 versus 2015-2019) were all < 0.001.

Table 2 shows the attributable mortality fractions and attributable deaths associated with PM_2.5_, PM_10_ and PM_2.5-10_ during March and May in 2020 and average values in 2015-2019. AFs and ADs were higher in 2020 than 2015-2019. We estimated that 10.21% (95% CI: 7.13, 13.31) of deaths were attributable to PM_2.5_ in first three months of pandemic in 2020, whereas the average AF was only 2.44 (95% CI: 1.63, 3.23) in 2015-2019, such disparity was consistent across all sex and age groups and was similar for PM_2.5-10_ and PM_10_. During the March to May, 20,062, 15,112, and 7,193 all-cause deaths were estimated to be attributable to PM_2.5_, PM_10_ and PM_2.5-10_, in 2020, approximately 5 times higher than the average values in 2015-2019.

**Table 2:**
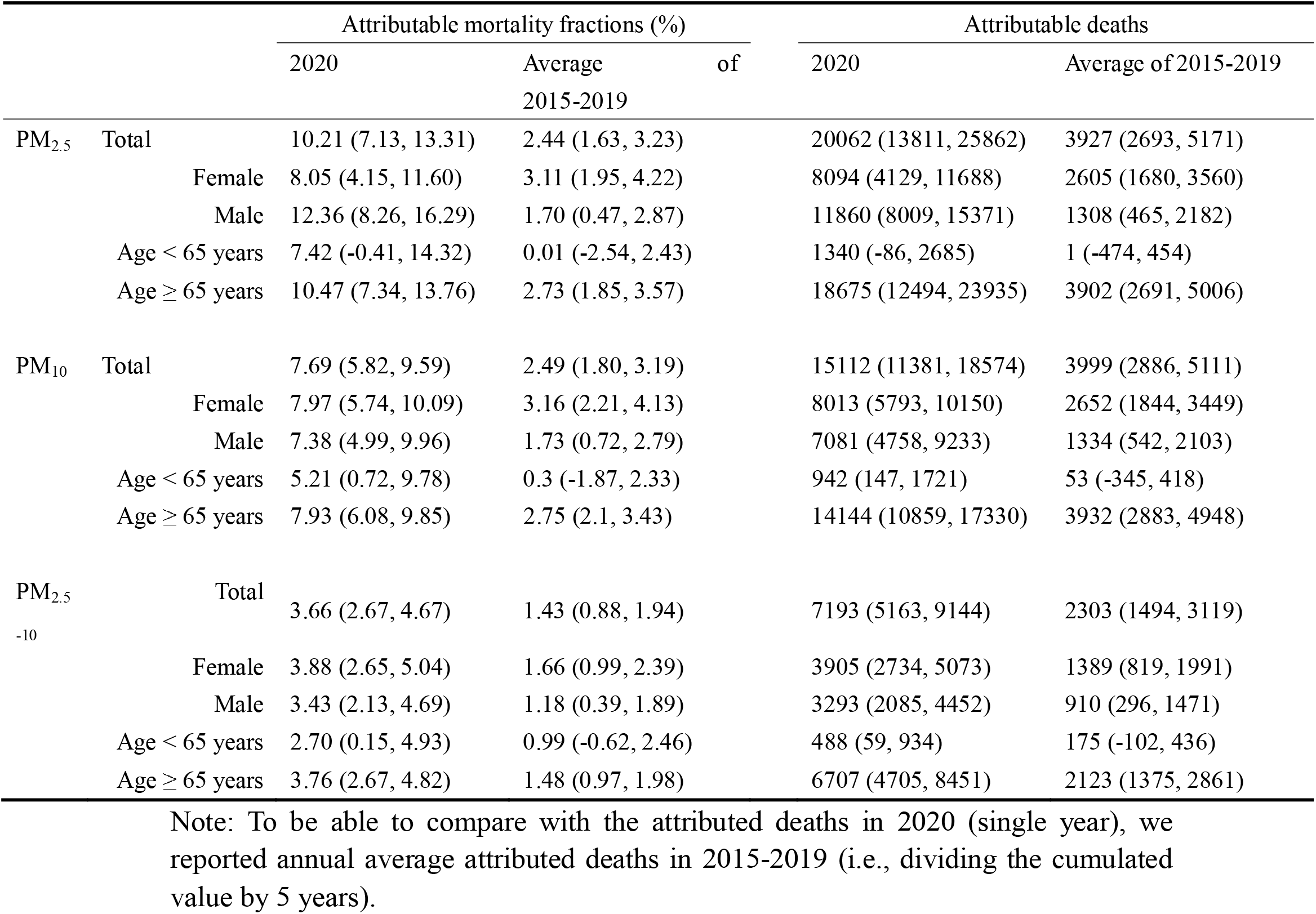
Attributable fractions and attributable counts of all-cause mortality due to PM_2.5_ and PM_10_ during the March to May in 2020 and 2015-2019, respectively.

Sensitivity results in Figure 3 consistently showed a stronger PM-mortality association during 2020 pandemic period compared to the associations during the same months in each year of 2015-2019, despite the variations after adjusting for different gaseous pollutants. Likewise, when we used different lag days of PM (lag 0-4 and lag 0-5 for PM_2.5_, lag 0-3 and lag 0-4 for PM_10_, lag 0-2 and lag 0-3 for PM_2.5-10_), the effects remained higher during 2020 pandemic period than the same period in 2015-2019 (Figure S2).

**Fig. 3:**
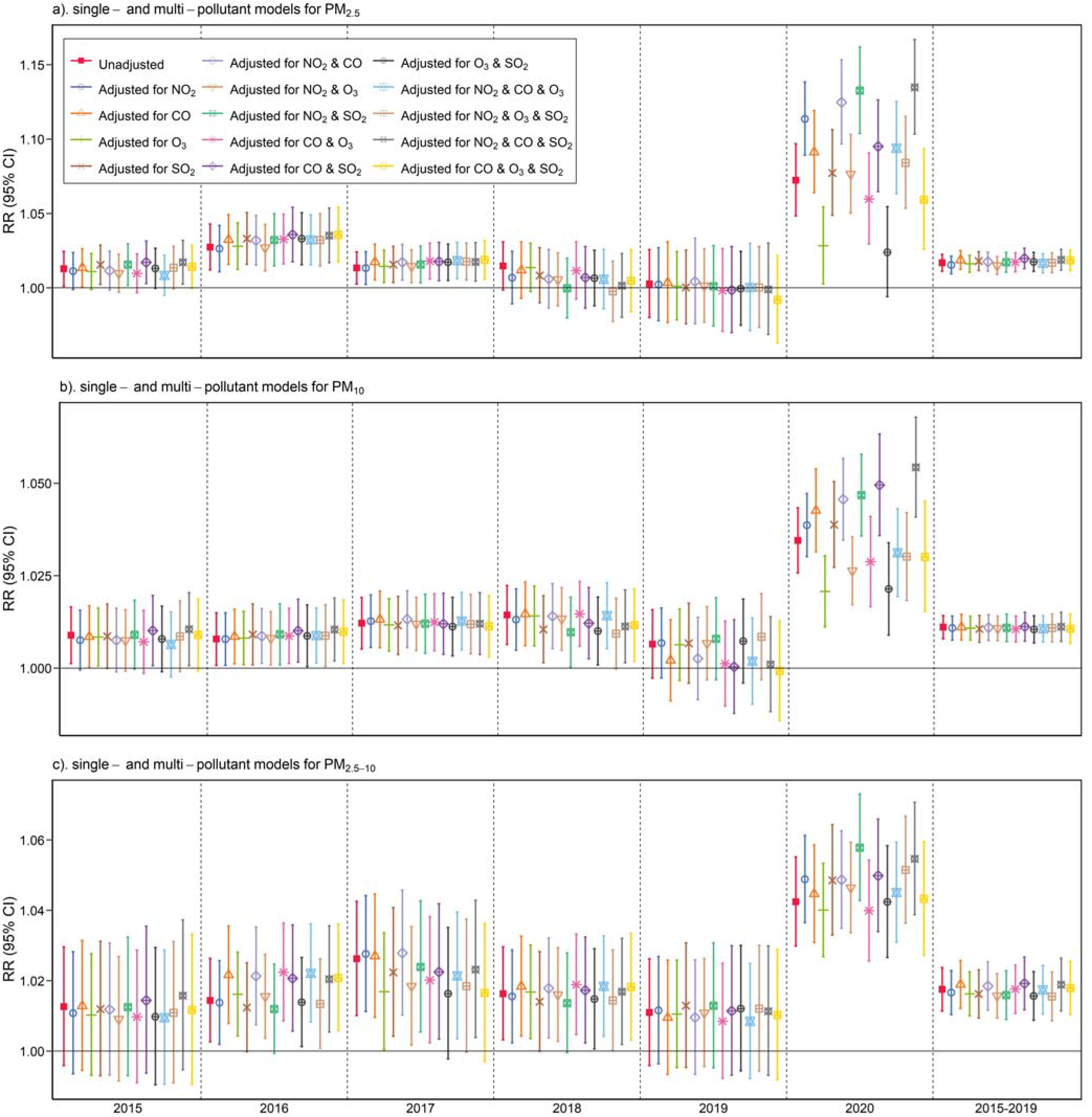
Cumulated RR (mean and 95%CI) of all-cause mortality risks associated with per 10 μg/m^3^ increase in PM concentrations in single- and multi-pollutant models.

## 4. Discussion

To our best knowledge, this the first study to investigate the relationship between PM air pollution and daily all-cause mortality during the COVID-19 pandemic period in the world. In this study, we examined the effects of PM (PM_2.5_, PM_10_, and PM_2.5-10_) on daily all-cause mortality in 107 Italian provinces and compared the mortality risks and mortality burdens associated with PM before and during the COVID-19 pandemic. The mortality risks (vulnerability) and burden associated with PM_2.5_, PM_10_ and PM_2.5-10_ in the pandemic were significantly higher than risks estimated in 2015-2019. People aged ≥ 65 years were consistently at higher risk than younger people in both pandemic period and normal periods.

Exposure to PM air pollution has been identified as the risk factor for excess mortality.^37 38^ Our results are in line with a previous multi-country epidemiological study,^39^ in which it observed a 0.65% (95% CI: 0.26%, 1.04%) increase in all-cause mortality risk per 10 μg/m^3^ increase at lag 0-1 of PM_10_ in 18 Italian cities during 2006-2015. Although the effect estimate is slightly lower than ours for PM_10_ at lag 0-2 [increased risk:1.11% (95% CI: 0.79%, 1.42%)], such difference in the PM_10_-mortality associations could be ascribed to the heterogeneity in period and season, as PM-mortality association might be stronger during cold months that we chose.^40^

The plausible pathways by which ambient PM air pollution may impact increased mortality risks have been well documented. Biologically, particles less than 10 μm in diameter^41^ have been acknowledged to be responsible for oxidative stress and chronic inflammation status that causes the hyper-activation of the immune system and the life-threatening respiratory disorders.^42 43^ The rapid and direct passage of smaller sized particles such as PM_2.5_ even escape into the blood stream and directly affect other organs far away from the lungs through systemic circulation.^44^ In addition, another plausible pathway for these adverse impacts is that PM may act as both a carrier and substrate of other toxic substance (e.g. carbonaceous elements,^45^ hazardous metals,^46^ etc.) and thus influence their fate and transport in the environment and reaching susceptible receptors.

In this study, we found that PM-attributed deaths during COVID-19 pandemic in 2020 were 5 times higher than the same months during 2015-2019, despite a lower PM level. One potential explanation is that PM may contribute to the COVID-19 related deaths. Most current studies have reported a positive association between ambient PM_2.5_ or PM_10_ and COVID-19 deaths,^27 47-53^ although they were limited by the inaccurate official reports of COVID-19 deaths. A review study highlighted the potential role of PM in the spread of COVID-19, focusing on Italian cities in which correspondence between poor air quality and COVID-19 induced mortality was the starkest yet.^54^ First, COVID-19 could have an air transmission^55 56^ and atmospheric PM could create a suitable environment for transporting the virus at greater distances than those considered for close contact.^57^ Second, PM has been shown to induce inflammation in lung cells^58^ and exposure to PM could increase the susceptibility and severity of the COVID-19 patient symptoms.^59^

However, the increase of PM-related mortality during pandemic period could also be from non-COVID-19 causes, particularly for those vulnerable populations (e.g., elderly people, people with chronic diseases). For example, during the early days of the COVID-19 outbreak, the regional disparities in health-care resource availability and accessibility^60^ could play an important role in the change of the vulnerability to PM related deaths. In March, though Italy’s health system has 3.2 hospital beds per 1000 people (as compared with 2.8 in the United States), it was over occupied to meet the needs of rapid surge of COVID-19 cases.^61^ The health needs created by the coronavirus pandemic went well beyond the capacity of national health system, diagnostic, therapeutic, and preventive interventions were scarce and rationed.^62 63^ In the worst scenario, patients with PM-related diseases would die while waiting for needed resources (e.g. ICU, ventilator and acute care).^64^ This hypothesis is supported by observations showing a significant decrease in hospitalization rates for acute coronary syndrome (13.3 admissions per day versus 18.0)^65^ and acute myocardial infarction (a reduction of 52.1% in North Italy, 59.3% in Central Italy, and 52.1% in South Italy)^66^ compared with the equivalent time in 2019 in Italy.

We also investigated the different effects between gender and age groups in the pandemic and normal periods. With regards to the age-stratified analysis, older people generally have a higher risk of PM-related death risk in both pandemic period and pre-pandemic periods, which was consistent with most previous studies.^67 68^ Though the detailed reasons are still unclear, pre-existing cardiovascular and respiratory diseases are more prevalent in elderly people, and it may enable the elderly to be more susceptible to ambient PM air pollution.

There are some limitations to this study. First, like most case-crossover or time-series studies, we used province-level air pollution to represent the individual-level exposure, which is likely to cause random exposure assessment error and thus underestimate the PM-mortality associations. However, since the same design was applied to the 2020 and 2015-2019 period, this error is not likely to affect our main findings. Second, we were not able to assess the actual indoor exposures to PM, which might be important because people would increase the indoor time during the lockdown period. Finally, we cannot exclude the COVID-19 deaths from our analyses because the daily data were not available at province level. Therefore, we were uncertain about whether the increase in PM-mortality association was due to COVID-19 deaths or deaths due to other causes.

In conclusion, with a large nationwide data set covering 107 Italian provinces, we observed significantly increased impacts of PM on all-cause mortality during the pandemic period compared to pre-COVID-19 periods. This suggests the historical exposure-response relationship between PM and mortality may underestimate the health impacts of PM during the COVID-19 pandemic, although air pollution concentrations declined. To date, the second wave of pandemic is ongoing in many Europe countries and no sign shows the end of the COVID-19 pandemic worldwide. Therefore, we would expect our findings to be helpful to guide future public health policy and environmental protection strategies such as making early warning systems for highly polluted regions, and giving close attention to vulnerable groups.

## Data Availability

Original data are open sources.

## Contributors

TY, RX, WY, YG, and SL designed the study. TY, RX, WY, and ZC collected data. TY did statistical analyses. TY and WY wrote the manuscript. RX, YG and SL provided scientific comments on the manuscript. All authors provided critical conceptual input, analyzed and interpreted data, and critically revised the report.

## References

1. Huang C, Wang Y, Li X, et al. Clinical features of patients infected with 2019 novel coronavirus in Wuhan, China. The Lancet 2020;395(10223):497–506. doi: 10.1016/s0140-6736(20)30183-5

2. Burnett R, Chen H, Szyszkowicz M, et al. Global estimates of mortality associated with long-term exposure to outdoor fine particulate matter. Proceedings of the National Academy of Sciences 2018;115(38):9592–97. doi: 10.1073/pnas.1803222115

3. Yang B-Y, Guo Y, Markevych I, et al. Association of Long-term Exposure to Ambient Air Pollutants With Risk Factors for Cardiovascular Disease in China. JAMA Network Open 2019;2(3):e190318–e18. doi: 10.1001/jamanetworkopen.2019.0318

4. Zheng H, Kong S, Chen N, et al. Significant changes in the chemical compositions and sources of PM2.5 in Wuhan since the city lockdown as COVID-19. Science of The Total Environment 2020;739:140000. doi: 10.1016/j.scitotenv.2020.140000

5. Chen K, Wang M, Huang C, et al. Air pollution reduction and mortality benefit during the COVID-19 outbreak in China. The Lancet Planetary Health 2020 doi: 10.1016/s2542-5196(20)30107-8

6. Otmani A, Benchrif A, Tahri M, et al. Impact of Covid-19 lockdown on PM10, SO2 and NO2 concentrations in Sale City (Morocco). Sci Total Environ 2020;735:139541. doi: 10.1016/j.scitotenv.2020.139541

7. Kanniah KD, Kamarul Zaman NAF, Kaskaoutis DG, et al. COVID-19’s impact on the atmospheric environment in the Southeast Asia region. Sci Total Environ 2020;736:139658. doi: 10.1016/j.scitotenv.2020.139658

8. Gautam S. The influence of COVID-19 on air quality in India: a boon or inutile. Bull Environ Contam Toxicol 2020;104(6):724–26. doi: 10.1007/s00128-020-02877-y

9. Nakada LYK, Urban RC. COVID-19 pandemic: Impacts on the air quality during the partial lockdown in Sao Paulo state, Brazil. Sci Total Environ 2020;730:139087. doi: 10.1016/j.scitotenv.2020.139087

10. Son J-Y, Fong KC, Heo S, et al. Reductions in mortality resulting from reduced air pollution levels due to COVID-19 mitigation measures. Science of The Total Environment 2020;744:141012. doi: 10.1016/j.scitotenv.2020.141012

11. Tobias A, Carnerero C, Reche C, et al. Changes in air quality during the lockdown in Barcelona (Spain) one month into the SARS-CoV-2 epidemic. Sci Total Environ 2020;726:138540. doi: 10.1016/j.scitotenv.2020.138540

12. Chen K, Wang M, Huang C, et al. Air pollution reduction and mortality benefit during the COVID-19 outbreak in China. The Lancet Planetary Health 2020;4(6):e210–e12. doi: 10.1016/s2542-5196(20)30107-8

13. Le Quéré C, Jackson RB, Jones MW, et al. Temporary reduction in daily global CO2 emissions during the COVID-19 forced confinement. Nat Clim Chang 2020;10(7):647–53. doi: 10.1038/s41558-020-0797-x

14. Boccia S, Ricciardi W, Ioannidis JPA. What Other Countries Can Learn From Italy During the COVID-19 Pandemic. JAMA Internal Medicine 2020;180(7):927–28. doi: 10.1001/jamainternmed.2020.1447

15. Alicandro G, Remuzzi G, La Vecchia C. Italy’s first wave of the COVID-19 pandemic has ended: no excess mortality in May, 2020. The Lancet 2020 doi: 10.1016/s0140-6736(20)31865-1

16. Collivignarelli MC, Abbà A, Bertanza G, et al. Lockdown for CoViD-2019 in Milan: What are the effects on air quality? The Science of the total environment 2020;732:139280. doi: 10.1016/j.scitotenv.2020.139280 [published Online First: 2020/05/14]

17. Chauhan A, Singh RP. Decline in PM(2.5) concentrations over major cities around the world associated with COVID-19. Environmental research 2020;187:109634. doi: 10.1016/j.envres.2020.109634 [published Online First: 2020/05/18]

18. Martelletti L, Martelletti P. Air Pollution and the Novel Covid-19 Disease: a Putative Disease Risk Factor. SN Comprehensive Clinical Medicine 2020;2(4):383–87. doi: 10.1007/s42399-020-00274-4

19. Adhikari A, Yin J. Short-Term Effects of Ambient Ozone, PM(2.5,) and Meteorological Factors on COVID-19 Confirmed Cases and Deaths in Queens, New York. International journal of environmental research and public health 2020;17(11):4047. doi: 10.3390/ijerph17114047

20. Bashir MF, Ma BJ Bilal, et al. Correlation between environmental pollution indicators and COVID-19 pandemic: A brief study in Californian context. Environmental research 2020;187:109652–52. doi: 10.1016/j.envres.2020.109652 [published Online First: 2020/05/13]

21. Liang D, Shi L, Zhao J, et al. Urban Air Pollution May Enhance COVID-19 Case-Fatality and Mortality Rates in the United States. The Innovation doi: 10.1016/j.xinn.2020.100047

22. Gill JR, DeJoseph ME. The Importance of Proper Death Certification During the COVID-19 Pandemic. JAMA 2020

23. Raymond E, Thieblemont C, Alran S, et al. Impact of the COVID-19 Outbreak on the Management of Patients with Cancer. Target Oncol 2020;15(3):249–59. doi: 10.1007/s11523-020-00721-1

24. Liang W, Guan W, Chen R, et al. Cancer patients in SARS-CoV-2 infection: a nationwide analysis in China. Lancet Oncol 2020;21(3):335–37. doi: 10.1016/s1470-2045(20)30096-6 [published Online First: 2020/02/19]

25. Rosenbaum L. The Untold Toll — The Pandemic’s Effects on Patients without Covid-19. New England Journal of Medicine 2020;382(24):2368–71. doi: 10.1056/NEJMms2009984

26. Zylke JW, Bauchner H. Mortality and morbidity: the measure of a pandemic. JAMA 2020

27. Coker ES, Cavalli L, Fabrizi E, et al. The Effects of Air Pollution on COVID-19 Related Mortality in Northern Italy. Environ Resour Econ (Dordr) 2020:1–24. doi: 10.1007/s10640-020-00486-1 [published Online First: 2020/08/25]

28. Scortichini M, Schneider dos Santos R, De, et al. Excess mortality during the COVID-19 outbreak in Italy: a two-stage interrupted time series analysis. medRxiv 2020:2020.07.22.20159632. doi: 10.1101/2020.07.22.20159632

29. Powell H, Krall JR, Wang Y, et al. Ambient Coarse Particulate Matter and Hospital Admissions in the Medicare Cohort Air Pollution Study, 1999-2010. Environmental health perspectives 2015;123(11):1152–8. doi: 10.1289/ehp.1408720 [published Online First: 2015/04/15]

30. Cai J. humidity: Calculate Water Vapor Measures from Temperature and Dew Point. 2019

31. Armstrong BG, Gasparrini A, Tobias A. Conditional Poisson models: a flexible alternative to conditional logistic case cross-over analysis. BMC medical research methodology 2014;14(1):122.

32. Gasparrini A, Guo Y, Hashizume M, et al. Mortality risk attributable to high and low ambient temperature: a multicountry observational study. The Lancet 2015;386(9991):369–75. doi: 10.1016/S0140-6736(14)62114-0

33. Lian T, Fu Y, Sun M, et al. Effect of temperature on accidental human mortality: A time-series analysis in Shenzhen, Guangdong Province in China. Scientific reports 2020;10(1):8410. doi: 10.1038/s41598-020-65344-y

34. Gasparrini A, Leone M. Attributable risk from distributed lag models. BMC Medical Research Methodology 2014;14(1):55. doi: 10.1186/1471-2288-14-55

35. Generalized nonlinear models in R: An overview of the gnm package [program], 2020.

36. Viechtbauer W. Conducting meta-analyses in R with the metafor package. Journal of Statistical Software 2010;36(3)

37. Guo Y, Li S, Tian Z, et al. The burden of air pollution on years of life lost in Beijing, China, 2004-08: retrospective regression analysis of daily deaths. BMJ: British Medical Journal 2013;347:f7139. doi: 10.1136/bmj.f7139

38. Dai L, Zanobetti A, Koutrakis P, et al. Associations of Fine Particulate Matter Species with Mortality in the United States: A Multicity Time-Series Analysis. Environmental health perspectives 2014;122(8):837–42. doi: 10.1289/ehp.1307568

39. Liu C, Chen R, Sera F, et al. Ambient Particulate Air Pollution and Daily Mortality in 652 Cities. New England Journal of Medicine 2019;381(8):705–15. doi: 10.1056/NEJMoa1817364

40. Zhang Y, Fang J, Mao F, et al. Age- and season-specific effects of ambient particles (PM(1), PM(2.5), and PM(10)) on daily emergency department visits among two Chinese metropolitan populations. Chemosphere 2020;246:125723. doi: 10.1016/j.chemosphere.2019.125723 [published Online First: 2019/12/31]

41. Kim K-H, Kabir E, Kabir S. A review on the human health impact of airborne particulate matter. Environment international 2015;74:136–43. doi: https://doi.org/10.1016/j.envint.2014.10.005

42. Shi Y, Wang Y, Shao C, et al. COVID-19 infection: the perspectives on immune responses. Cell Death & Differentiation 2020;27(5):1451–54. doi: 10.1038/s41418-020-0530-3

43. Osornio-Vargas AR, Serrano J, Rojas-Bracho L, et al. In vitro biological effects of airborne PM2.5 and PM10 from a semi-desert city on the Mexico–US border. Chemosphere 2011;83(4):618–26. doi: https://doi.org/10.1016/j.chemosphere.2010.11.073

44. Nemmar A, Vanbilloen H, Hoylaerts MF, et al. Passage of Intratracheally Instilled Ultrafine Particles from the Lung into the Systemic Circulation in Hamster. American Journal of Respiratory and Critical Care Medicine 2001;164(9):1665–68. doi: 10.1164/ajrccm.164.9.2101036

45. Sharma SK, Mandal TK, Sharma A, et al. Carbonaceous Species of PM(2.5) in Megacity Delhi, India During 2012-2016. Bull Environ Contam Toxicol 2018;100(5):695–701. doi: 10.1007/s00128-018-2313-9 [published Online First: 2018/03/09]

46. Luo X, Zhao Z, Xie J, et al. Pulmonary bioaccessibility of trace metals in PM(2.5) from different megacities simulated by lung fluid extraction and DGT method. Chemosphere 2019;218:915–21. doi: 10.1016/j.chemosphere.2018.11.079 [published Online First: 2019/01/06]

47. Zhu Y, Xie J, Huang F, et al. Association between short-term exposure to air pollution and COVID-19 infection: Evidence from China. Science of The Total Environment 2020;727:138704. doi: https://doi.org/10.1016/j.scitotenv.2020.138704

48. Fattorini D, Regoli F. Role of the chronic air pollution levels in the Covid-19 outbreak risk in Italy. Environmental pollution 2020;264:114732. doi: 10.1016/j.envpol.2020.114732 [published Online First: 2020/05/11]

49. Cole MA, Ozgen C, Strobl E. Air Pollution Exposure and Covid-19 in Dutch Municipalities. Environ Resour Econ (Dordr) 2020:1-30. doi: 10.1007/s10640-020-00491-4 [published Online First: 2020/08/25]

50. Wu X, Nethery RC, Sabath BM, et al. Exposure to air pollution and COVID-19 mortality in the United States: A nationwide cross-sectional study. medRxiv 2020:2020.04.05.20054502. doi: 10.1101/2020.04.05.20054502

51. Vasquez-Apestegui V, Parras-Garrido E, Tapia V, et al. Association Between Air Pollution in Lima and the High Incidence of COVID-19: Findings from a Post Hoc Analysis. Res Sq 2020 doi: 10.21203/rs.3.rs-39404/v1 [published Online First: 2020/07/24]

52. Jiang Y, Wu XJ, Guan YJ. Effect of ambient air pollutants and meteorological variables on COVID-19 incidence. Infect Control Hosp Epidemiol 2020;41(9):1011–15. doi: 10.1017/ice.2020.222 [published Online First: 2020/05/12]

53. Yao Y, Pan J, Wang W, et al. Association of particulate matter pollution and case fatality rate of COVID-19 in 49 Chinese cities. The Science of the total environment 2020;741:140396. doi: 10.1016/j.scitotenv.2020.140396 [published Online First: 2020/06/28]

54. Comunian S, Dongo D, Milani C, et al. Air Pollution and Covid-19: The Role of Particulate Matter in the Spread and Increase of Covid-19’s Morbidity and Mortality. International journal of environmental research and public health 2020;17(12) doi: 10.3390/ijerph17124487 [published Online First: 2020/06/26]

55. Zhou F, Yu T, Du R, et al. Clinical course and risk factors for mortality of adult inpatients with COVID-19 in Wuhan, China: a retrospective cohort study. The Lancet 2020;395(10229):1054–62. doi: 10.1016/S0140-6736(20)30566-3

56. van Doremalen N, Bushmaker T, Morris DH, et al. Aerosol and Surface Stability of SARS-CoV-2 as Compared with SARS-CoV-1. New England Journal of Medicine 2020;382(16):1564–67. doi: 10.1056/NEJMc2004973

57. Wei M, Liu H, Chen J, et al. Effects of aerosol pollution on PM2.5-associated bacteria in typical inland and coastal cities of northern China during the winter heating season. Environmental pollution 2020;262:114188. doi: https://doi.org/10.1016/j.envpol.2020.114188

58. Farina F, Sancini G, Battaglia C, et al. Milano Summer Particulate Matter (PM10) Triggers Lung Inflammation and Extra Pulmonary Adverse Events in Mice. PloS one 2013;8(2):e56636. doi: 10.1371/journal.pone.0056636

59. Li B, Yang J, Zhao F, et al. Prevalence and impact of cardiovascular metabolic diseases on COVID-19 in China. Clinical Research in Cardiology 2020;109(5):531–38. doi: 10.1007/s00392-020-01626-9

60. Ji Y, Ma Z, Peppelenbosch MP, et al. Potential association between COVID-19 mortality and health-care resource availability. Lancet Glob Health 2020;8(4):e480. doi: 10.1016/s2214-109x(20)30068-1 [published Online First: 2020/02/29]

61. Rosenbaum L. Facing Covid-19 in Italy - Ethics, Logistics, and Therapeutics on the Epidemic’s Front Line. N Engl J Med 2020;382(20):1873–75. doi: 10.1056/NEJMp2005492 [published Online First: 2020/03/19]

62. Ji Y, Ma Z, Peppelenbosch MP, et al. Potential association between COVID-19 mortality and health-care resource availability. The Lancet Global Health 2020;8(4):e480.

63. Emanuel EJ, Persad G, Upshur R, et al. Fair Allocation of Scarce Medical Resources in the Time of Covid-19. New England Journal of Medicine 2020;382(21):2049–55. doi: 10.1056/NEJMsb2005114

64. Barrett K, Khan YA, Mac S, et al. Estimation of COVID-19-induced depletion of hospital resources in Ontario, Canada. Cmaj 2020;192(24):E640–e46. doi: 10.1503/cmaj.200715 [published Online First: 2020/05/16]

65. De Filippo O, D’Ascenzo F, Angelini F, et al. Reduced rate of hospital admissions for ACS during Covid-19 outbreak in Northern Italy. New England Journal of Medicine 2020

66. De Rosa S, Spaccarotella C, Basso C, et al. Reduction of hospitalizations for myocardial infarction in Italy in the COVID-19 era. European heart journal 2020;41(22):2083–88.

67. Kang SJ, Jung SI. Age-Related Morbidity and Mortality among Patients with COVID-19. Infect Chemother 2020;52(2):154–64. doi: 10.3947/ic.2020.52.2.154 [published Online First: 2020/06/12]

68. Han F, Yang X, Xu D, et al. Association between outdoor PM(2.5) and prevalence of COPD: a systematic review and meta-analysis. Postgrad Med J 2019;95(1129):612–18. doi: 10.1136/postgradmedj-2019-136675 [published Online First: 2019/09/09]

