## Supplementary Figure S1, and Figure S2 for "Vulnerability and burden of all-cause mortality associated with particulate air pollution increased during COVID-19 pandemic: a nationwide observed study in Italy"

**Online-only Supplements**

**Table of contents**

- **Figure S1.** RRs on different lag days of PM_2.5_, PM_2.5_ and PM_2.5-10_.
- **Figure S2.** Cumulative RRs on different lag days of PM_2.5_, PM_2.5_ and PM_2.5-10_.

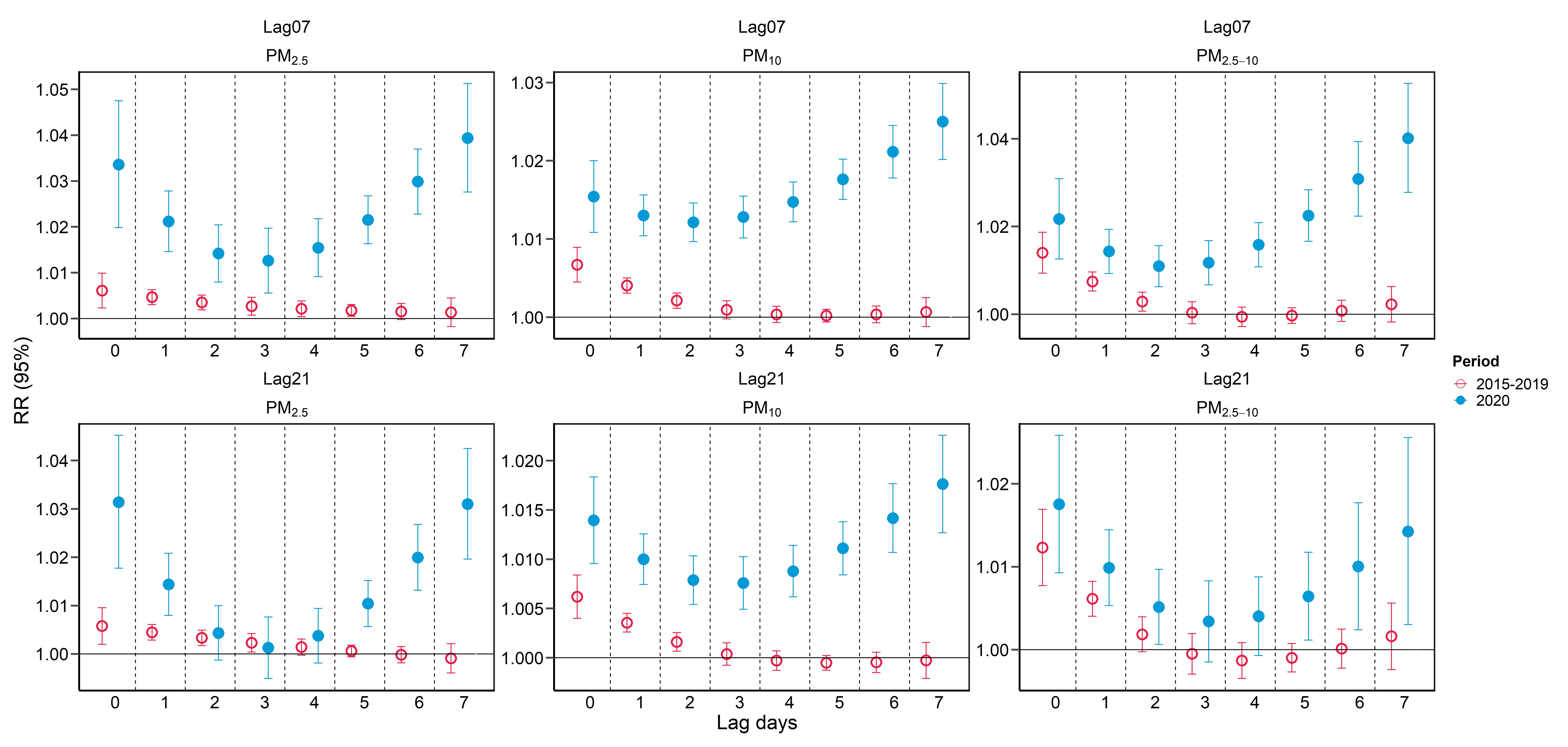

Figure S1. RRs on different lag days of PM_2.5_, PM_2.5_ and PM_2.5-10_.

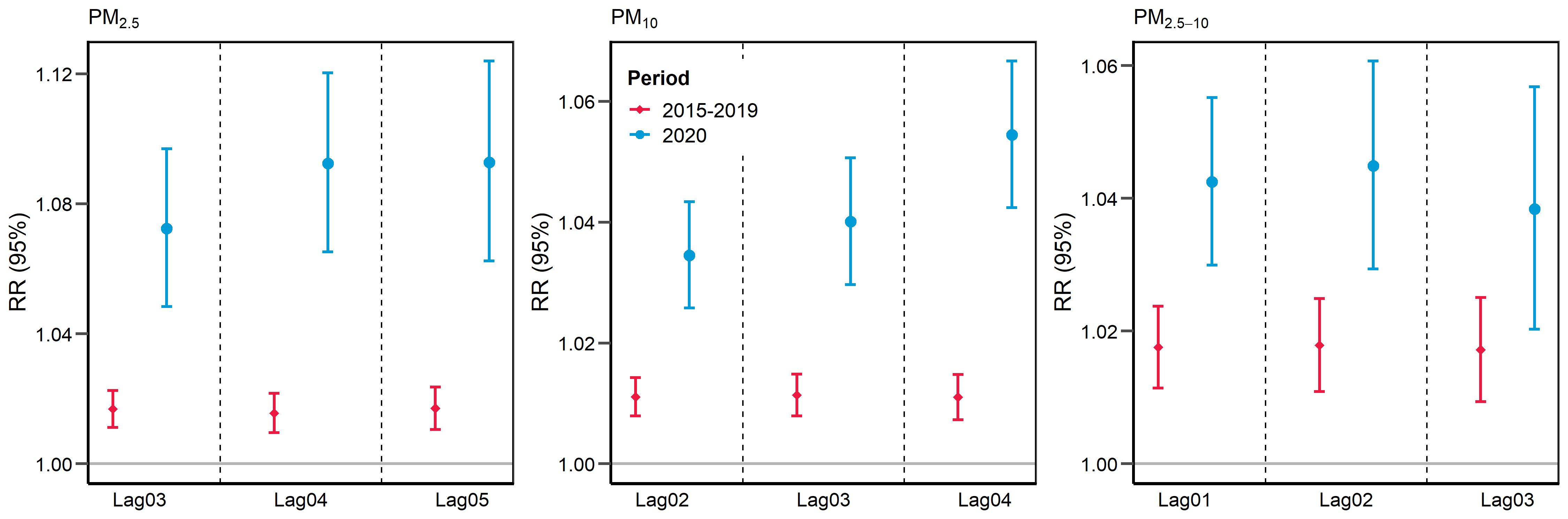

Figure S2. Cumulative RRs on different lag days of PM_2.5_, PM_2.5_ and PM_2.5-10_.
